# The Healthy Minds, Thriving Kids Project – a mental health and wellbeing prevention program for young people, results of a survey to understand the perspective of educators on program relevance and potential impact

**DOI:** 10.1101/2024.06.01.24308320

**Authors:** David Anderson, Jeffrey Chapman, Janine Domingues, Gabriella Bobadilla, Mimi Corcoran, Harold Koplewicz

## Abstract

**Background:** Healthy Minds Thriving Kids (HMTK) is a free to user mental health prevention program developed by the Child Mind Institute with the aim to normalize conversations about mental health and provide educators with wellness tools.

**Methods:** The HMTK program was available to view by registrants between 26^th^ January 2022 and 7^th^ September 2022 in the State of California. Participants viewed an introductory video for the program and a minimum of two skills videos before participating in an online survey.

**Results:** Of 68,861 registrants to the website 64,376 provided survey data. Post-pandemic levels of stress and anxiety were increased and 89.5% of respondents said young people required a greater degree of support than previously. Almost all educators (90%) want additional mental health and wellbeing tools for students and following review of HMTK >80% of respondents said they would use the program in their classrooms. Most (86.6%) found the program engaging and 85.1% found the program relevant to, and representative of, their student cohorts. More than three quarters (79.6%) said their students would find the program engaging and beneficial, and 18.8% more educators believed that HMTK was indicative of commitment to educators by the State of California in supporting students’ social and emotional learning.

**Conclusion:** This survey demonstrates that the HMTK program is a valuable and complementary resource to school curricula to improve the mental health and wellbeing of young people. It provides an easy to implement framework that school districts and administrators can allocate within their curriculum planning.

## Introduction

Childhood and adolescence presents a time of immense change for the developing brain [1] and is associated with increases in risky behavior, expression of strong emotions, and impulsivity. [2] Adolescents with mental health conditions are particularly vulnerable to social exclusion, discrimination, and stigma, which affect their readiness to seek help, leading to behavioral or educational difficulties, risk-taking behaviors and physical ill-health. [3] Mental health among children and adolescents in the USA and Europe has been declining for some time [4, 5] and depression, anxiety, and behavioral/conduct problems are prevalent and increasing. The extent of mental health issues among young people is well-validated, by the age of 24 the lifetime prevalence of mental health disorders is 75% [6], with 50% in place by the age of 14 years.

In the context of this landscape of pre-existing mental health decline, the SARS-CoV2 (COVID-19) pandemic has had a significant impact on mental wellbeing in both young people and adults, increasing reports of stress, anxiety, and depression by up to 25%. [7–13] The effects of COVID-19 on young people’s mental health has been varied, impacting readiness to learn and a range of other factors. [9, 14–20] These factors include societal disruption and separation from friends and family, with a pronounced impact on finances, housing, social support, relationships and daily routines. Stay-at-home orders led to pandemic-related job losses or a loss of income until a return to work was possible, increasing instability experienced by young people and their families. Throughout the pandemic, worry about the lethal consequences of COVID-19 among young people increased social isolation. Similarly, grief and trauma due to loss of a family member or caregiver due to COVID-19 will have a life-long effect on the thousands of young people who experiences these events. Post-COVID, it is clear across many countries that increases in abuse, neglect, violence or discrimination have occurred as a result of COVID-19. It is suggested this may be as a result of social isolation, lack of interaction with structural support services such as child welfare, and caregivers becoming overwhelmed by stressors in their lives.

Whilst addressing deteriorating mental health and wellbeing is considered a priority for intervention, treatment gaps remain in the USA, particularly for anxiety and behavioral problems. [21] Gaps may be apparent for range of reasons including, but not limited to, fair and equitable reimbursement from insurers for behavioral health treatment, lack of funding particularly within the education system to explore and optimize health literacy, demand for child and adolescent psychiatric services outstripping the supply available, and a need to integrate child and adolescent mental health and well-being services within the wider landscape of health, familial, educational, or legal contexts. Improving mental health and wellbeing among children and adolescents must be a national and global priority from an early age and has led to the development of the Healthy Minds, Thriving Kids (HMTK) program.

HMTK is an innovative mental health promotion program developed by the Child Mind Institute, a not-for-profit organization dedicated to supporting children and their families with mental health and learning disorders. Educators are confronted with a fast-growing number of young people experiencing mental health distress, which has been exacerbated by the pandemic. With mental health services for young people under strain there is increasing pressure on schools and educators to support young people. Across the USA, educators report that they do not receive sufficient training to be able to provide mental health support to their students, a fact that has been established by Mental Health America exhorting strategy development at local, state and national level (www.mhanational.org). The HMTK program was developed to provide educators with a child and adolescent mental health skill-building framework that would complement any existing social and emotional learning programs while also teaching unique, evidence-based skills. The development of HMTK was predicated on the belief that taking a coordinated and evidence-based approach to mental health and wellbeing in children and adolescents in school can improve their outcomes.

Specific aims of HMTK are to normalize conversations about mental health and wellbeing and provide educators with wellness tools. The program is free of charge to end users to teach skills that children and adolescents can use throughout their whole lives. Program material is based on well-established concepts and techniques in cognitive behavioral therapy (CBT). CBT is well established as a treatment for a wide range of mental health conditions in young people. [22] It is commonly used to treat anxiety and depression and can be useful for other mental health problems. It is considered the gold-standard evidence-based approach in mental health treatment, based on the concept that the interconnected nature of thoughts, feelings, physical sensations, and actions allows patients to identify and modify specific patterns of thinking and behavior in order to improve emotional health. CBT is skills-focused and data-driven, equipping individuals with tools to address problems and alleviate symptoms. Through CBT, young people gain skills to understand and cope with distressing emotions and to engage in more positive interactions with family and peers. [23] School-based CBT programs have shown reductions in symptoms of depression and anxiety as well as improved coping skills. [23]

CBT-based topics covered in HMTK include understanding feelings, relaxation skills, understanding thoughts and their impact on feelings and behaviors, management of intense emotions, and mindfulness. The program is available for three school age ranges – elementary, middle, and high school – and in two languages (Spanish and English). All HMTK videos, implementation guides, and worksheets can be accessed via www.childmind.org/healthyminds.

All HMTK project resources and content were developed by clinicians, educators, and staff of the Child Mind Institute, with video resource creation coordinated by the production company M SS NG P ECES. With the support of the State of California, Governor Gavin Newsom and First Partner Jennifer Siebel Newsom, and the California State Legislature, the program was piloted among Californian educators. A survey across California of educators registering to access the HMTK video resources was undertaken to explore the issues they face at an advanced phase of the pandemic, their general perception of mental health and wellbeing resources for educators, and the potential applicability and impact of the HMTK project for their students.

## Materials and methods

### Survey development

The survey instrument was developed in consultation with a range of stakeholders, including the authors as well as Lake Research Partners, and it was implemented by the Performance Development Group (PDG) and InsiteHub. All respondents were educators in the State of California, enrolled by PDG via a specifically designed educator portal to view the HMTK program materials. All survey questions were developed using a [5-point](https://brandfish.sharepoint.com/:x:/s/BrandfishClients/EQCv3mDhvYpJtR1eqLo2VSMB9FWUnAHltc5T3v6Vu673fg) Likert scale from Strongly Disagree to Strongly Agree. The questions and top-line data can be found in Supporting Information.

### Survey design

Participants accessed the HMTK portal to view HMTK video materials between 26^th^ January 2022 and 7^th^ September 2022. On entry to the portal, participants consented to participate and completed a pre-program survey, viewed an introductory video of the program and viewed at least two of the five skills videos. Following review of HMTK materials, registrants participated in a follow up survey to explore their perceptions of the materials and their utility in the classroom. All registrants of the HMTK portal completing the pre-program and post-program surveys, developed by CMI and administered by PDG, were eligible to receive a financial reward of $100. The HMTK portal was closed at the end of the survey period. The survey data were analysed using contingency tables that were created using QPSMR version CL 64 2021.2c and data analysed in Excel (see Supporting Information).

### Ethics and consent

Whilst no industry statutes exist in the USA for the conducting of market research, Lake Research Partners (LRP) are members of the American Association for Public Opinion Research (AAPOR). All questionnaires were developed by LRP and CMI to be compliant with and adherent to the AAPOR Code of Professional Ethics and Practices, which is comparable to counterpart guidelines in Europe. Informed consent was provided via self-selected participation in adherence with AAPOR guidance. All data were further aggregated to retain complete anonymization, and all personal data was collected under AAPOR standards. Generated data only assessed and documented feedback on the materials of the HMTK program and did not include generalisable knowledge including biomedical or behavioral research or school records. As such, this survey does not meet criteria found in 45 CFR 46.104(d) and as such is exempt from IRB oversight and IRB exemption was granted on 15^th^ May 2024 (Pro00079335, Advarra IRB registered with OHRP and FDA under IRB#00000971).

## Results

Between 26^th^ January 2022 and 7^th^ September 2022, 72,680 people registered with the HMTK portal to review materials (**Table 1**). Of these, 72,097 are English speakers and 583 Spanish speakers. Participants represented 8,443 schools and 990 school districts, and 60,003 registrants fulfilled their financial incentive (82.5%). Original data can be found in the Supporting information.

**Table 1.** Overview of registrants to the HMTK portal as of 24/10/2022.

| Parameter | Number of respondents n(%) |
| --- | --- |
| Total registrants | 72680 |
| Schools represented (n) | 8443 |
| School districts represented (n) | 990 |
| Number of Spanish-speaking registrants | 583 (0.8%) |
| Number of English-speaking registrants | 72097 (99.2%) |
| Educational setting |  |
| Kindergarten to Grade 5 (K–5) | 40552 (55.8%) |
| Grades 6–8 | 18672 (25.7%) |
| Grades 9–12 | 13456 (18.5%) |

Of registrants to the website, 68,861 opted to take part in the survey to evaluate the impact of COVID-19 on the mental health and wellbeing of young people, and up to 64,376 provided data for a survey following their review of HMTK video materials. Almost half of all registrants accessed the program on a desktop computer (48.9%), with 50.6% accessing content on their smartphone and 0.5% accessing via a tablet. Most registrants were Kindergarten to Grade 5 educators (K–5) n=40552 (55.8%), 25.7% (n=18,672) were teaching Grades 6–8, and 18.5% (n=13,456) were teaching Grades 9–12.

Evaluation of the impact of COVID-19 on mental health and wellbeing on young people demonstrated a high agreement (83.3%) that COVID-19 has heightened stress and anxiety among young people (**Fig. 1**) and results were consistent regardless of the student cohort (**Table 2**). Similarly, educators indicated that rates of disruptive behavior in the classroom were consistently increased post-COVID-19 across all student age groups. Most educators responded that young people required greater support with respect to social and emotional learning following the pandemic compared with before (89.5%) (**Fig 2**; **Table 3**). Most educators said they would benefit from having more social and emotional learning resources for their students (90.2%).

**Fig 1.**
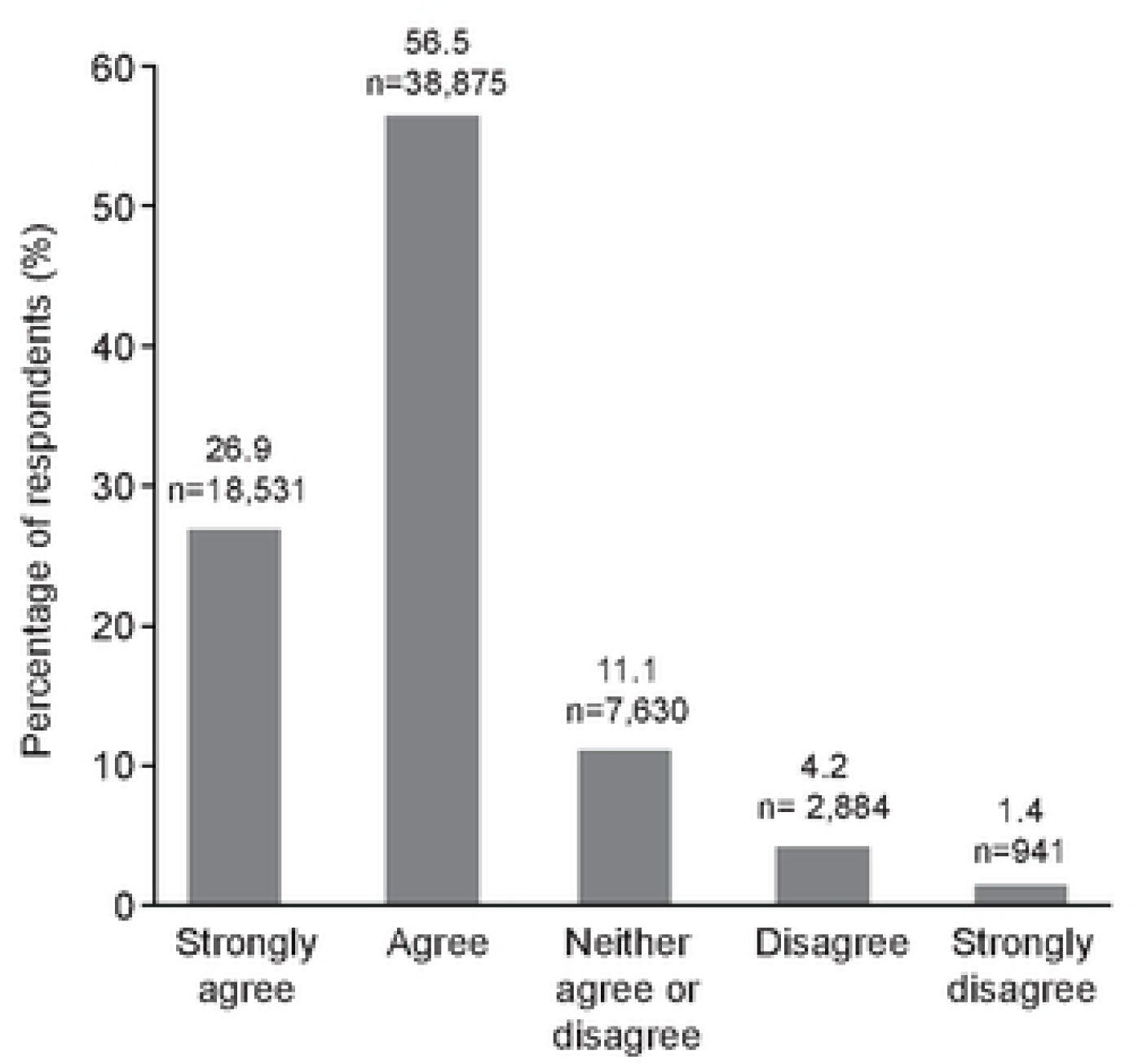
Impact of COVID-19 on mental health and wellbeing. **Q. Since Covid, my students are demonstrating more signs of stress or anxiety in school.**

**Fig 2.**
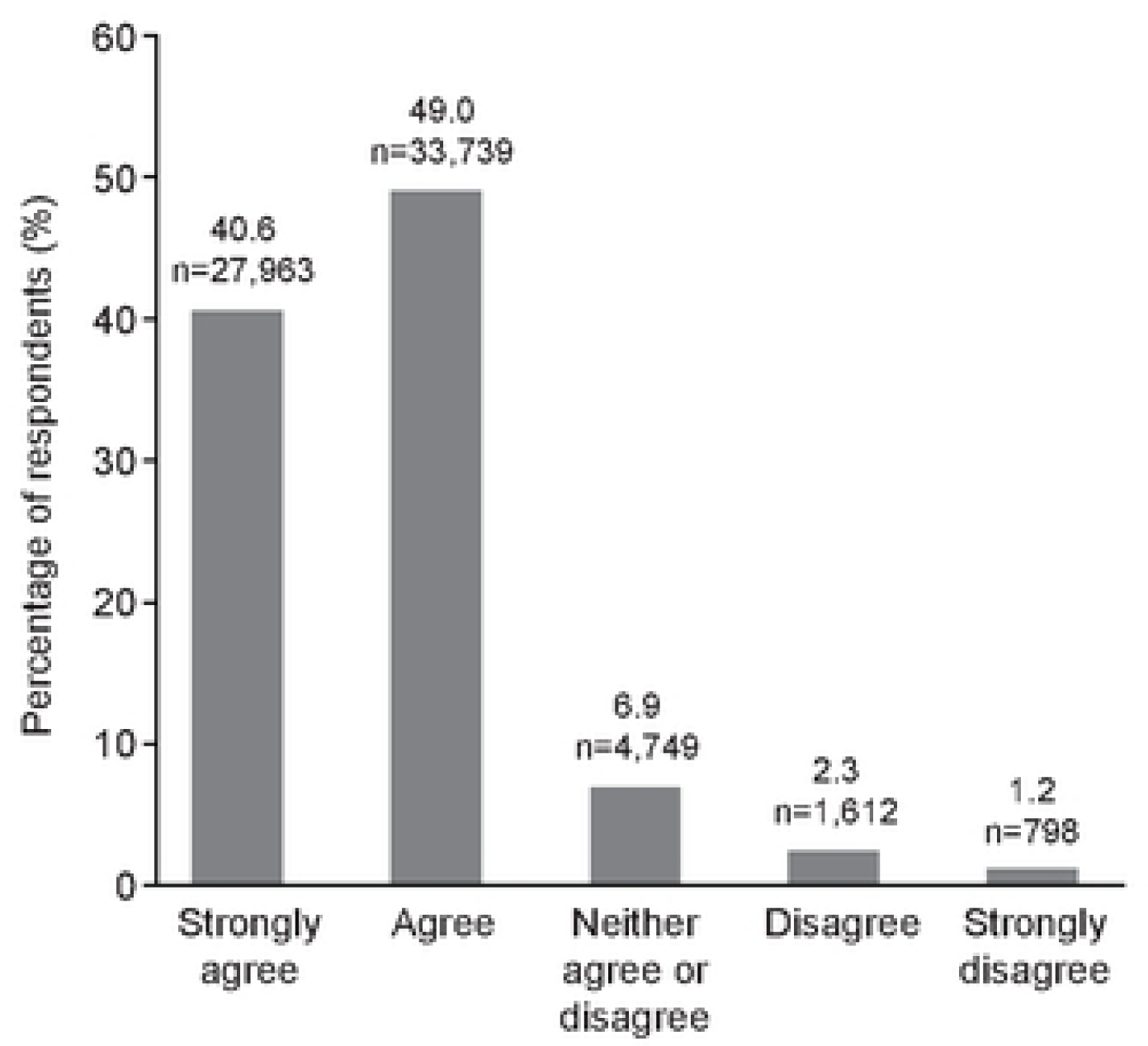
Requirement for increased emotional and social support from educators post-COVID-19. **Q. Since Covid, I am having to support more of my students’ social and emotional learning.**

**Table 2.** Impact of COVID-19 on specific educational cohorts. Base n=68,861.

|  | Strongly agree N(%) | Agree N(%) | Neither agree or disagree N(%) | Disagree N(%) | Strongly disagree N(%) |
| --- | --- | --- | --- | --- | --- |
| K–5 (n=37,478) | 8650 (23.1) | 21829 (58.2) | 4729 (12.6) | 1798 (4.8) | 472 (1.3) |
| Grade 6–8 (n=12,609) | 3884 (30.8) | 7049 (55.9) | 1116 (8.9) | 408 (3.2) | 152 (1.2) |
| Grade 9–12 (n=18,774) | 5997 (31.9) | 9997 (53.2) | 1785 (9.5) | 678 (3.6) | 317 (1.7) |

Q. Since Covid, my students are demonstrating more signs of stress or anxiety in school.

**Table 3.** Requirement for support in specific educational cohorts. Base n=68,861.

|  | Strongly agree N(%) | Agree N(%) | Neither agree or disagree N(%) | Disagree N(%) | Strongly disagree N(%) |
| --- | --- | --- | --- | --- | --- |
| K–5 (n=37,478) | 15,817 (42.2) | 18,169 (48.5) | 2,256 (6.0) | 815 (2.2) | 421 (1.1) |
| Grade 6–8 (n=12,609) | 4,629 (42.1) | 6,141 (48.7) | 792 (6.3) | 241 (1.9) | 129 (1.0) |
| Grade 9–12 (n=18,774) | 4,904 (36.4) | 9,429 (50.2) | 1,701 (9.1) | 556 (3.0) | 248 (1.3) |

Q. Since Covid, I am having to support more of my students' social and emotional learning.

Before review of the HMTK video program, almost a quarter of respondents felt that programs to support mental health and wellbeing were not presented in an engaging way, and this was particularly noted in educators teaching Grades 6–8 (19.2%) and Grades 9–12 (17.0%) compared with K–5 (9.4%). Following review of the video materials through the portal, almost all respondents agreed that students would benefit from engaging with the videos within the HMTK program (90.2%), and 79.6% of respondents said they thought their students would find the videos interesting and engaging (**Fig 3a,b**). More than 80% of educators said the videos provided insights into how to approach students’ social and emotional needs (87.3% (**Fig 4a**). Educators themselves found the videos engaging (86.6%) and were emotionally moved by the video content (65.7%), and 85.1% felt that the videos were representative of their student cohorts (**Fig 4b**).

**Fig 3.**
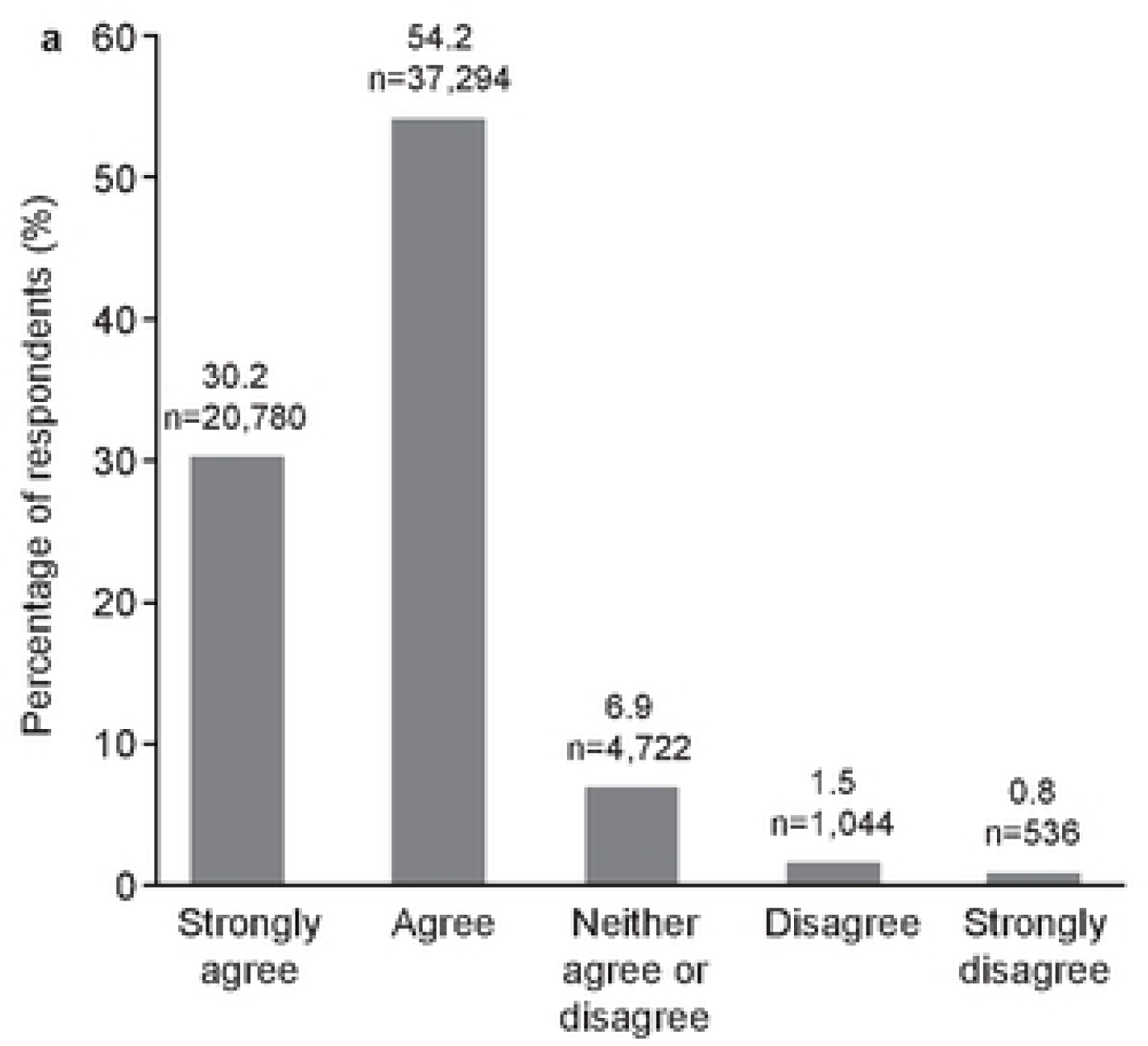
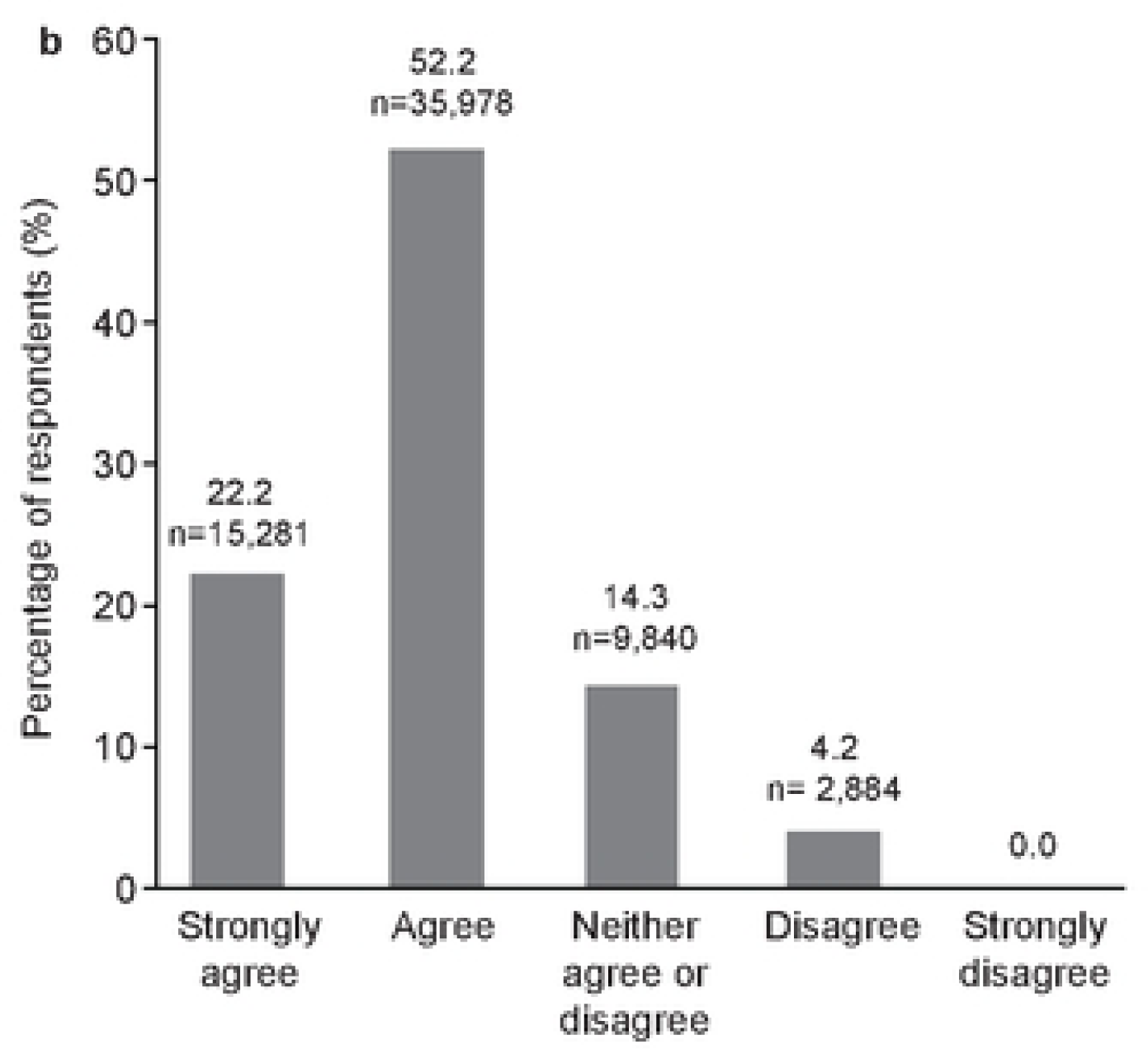
Perception of HMTK by students. a) my students would benefit from seeing the videos in this program; b) my students would find these videos interesting to watch. **a)** **Q. My students would benefit from seeing the videos in this program.** **b)** **R. My students would benefit from seeing the videos in this program.**

**Fig 4.**
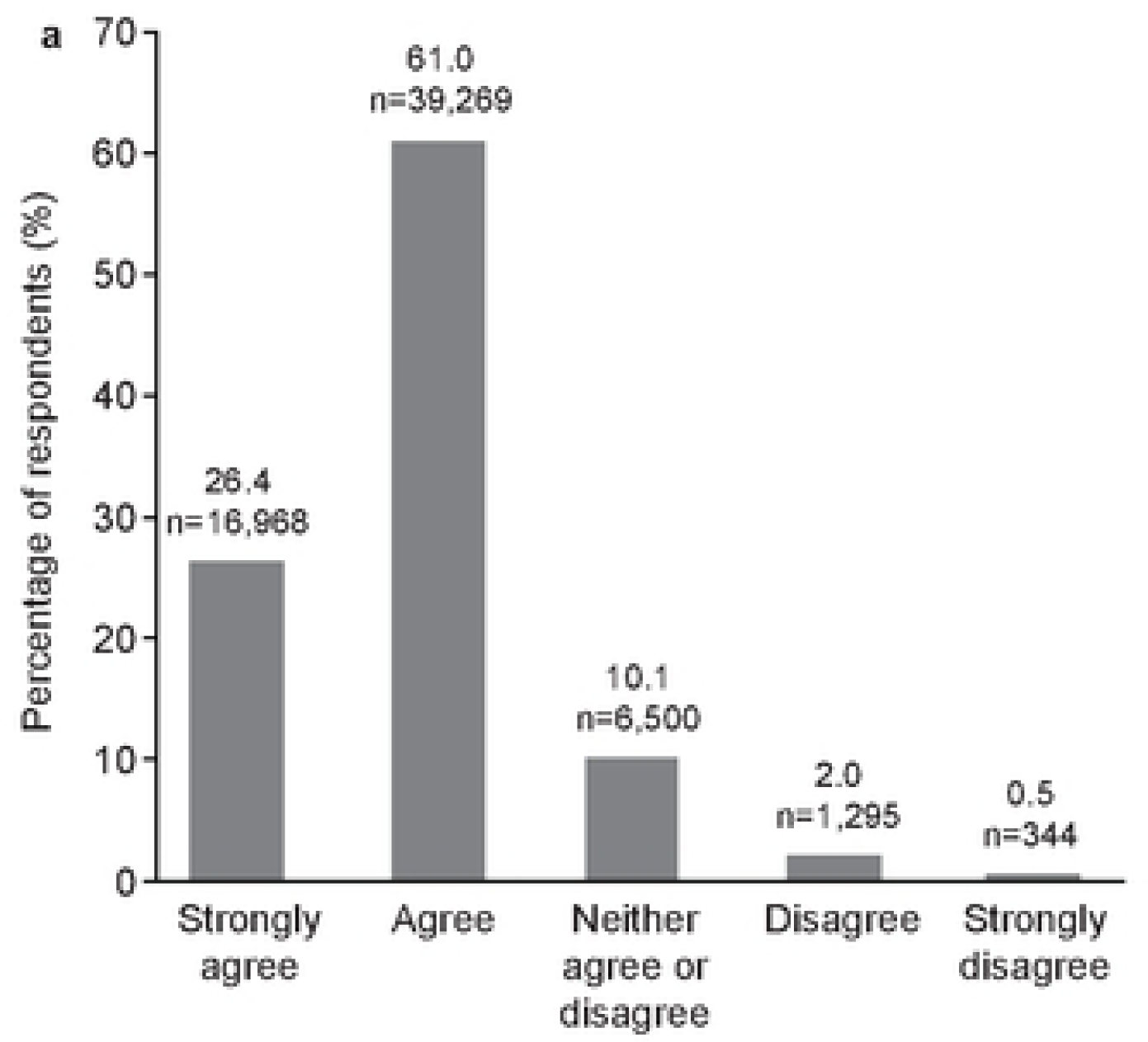
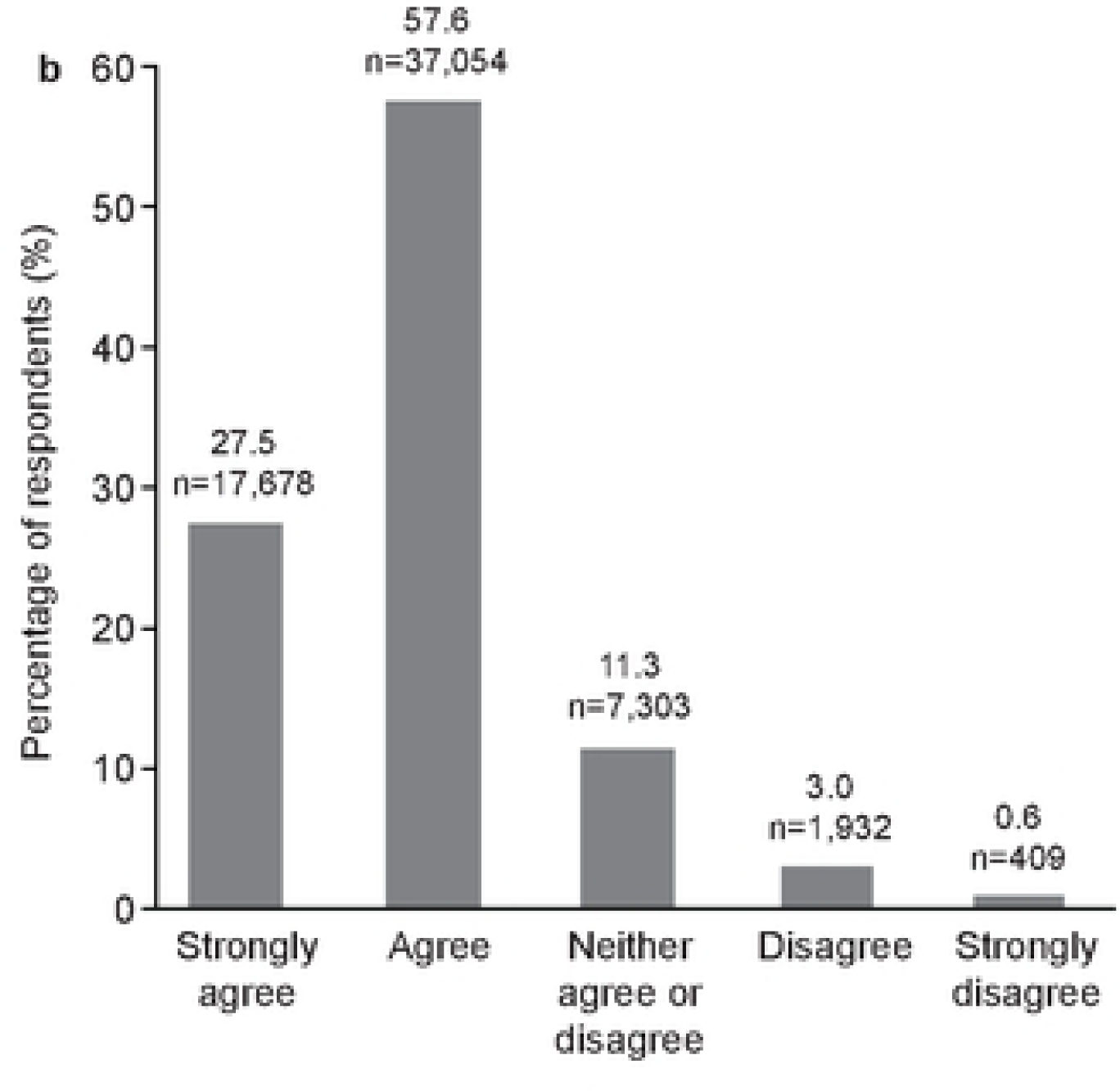
Educator perspectives of the HMTK program. a) Educator insights into how to approach students’ social and emotional needs; b) educators perspectives of HTMK materials. a) **Q. I gained insights on how to approach my students’ social and emotional learning needs watching these videos.** **b)** **Q. These videos did a good job representing the diversity of California school students.**

More than 80% of educators said they were likely to use the videos in their classroom mental health and wellbeing programs (82.2%) (**Fig 5**). When asked if educators believed the State of California was committed to supporting the social and emotional learning of its students, there was a striking increase in agreement between pre- and post-surveys. Before engaging with the HMTK program, only a quarter of educators felt the State of California was committed to supporting students’ social and emotional learning. Following their review of the HMTK video materials, educators demonstrated an 18.8% increase in their belief that the state of California was committed to resources supporting students’ mental health and wellbeing, and only 10% felt that the State of California was not committed (**Fig 6**).

**Fig 5.**
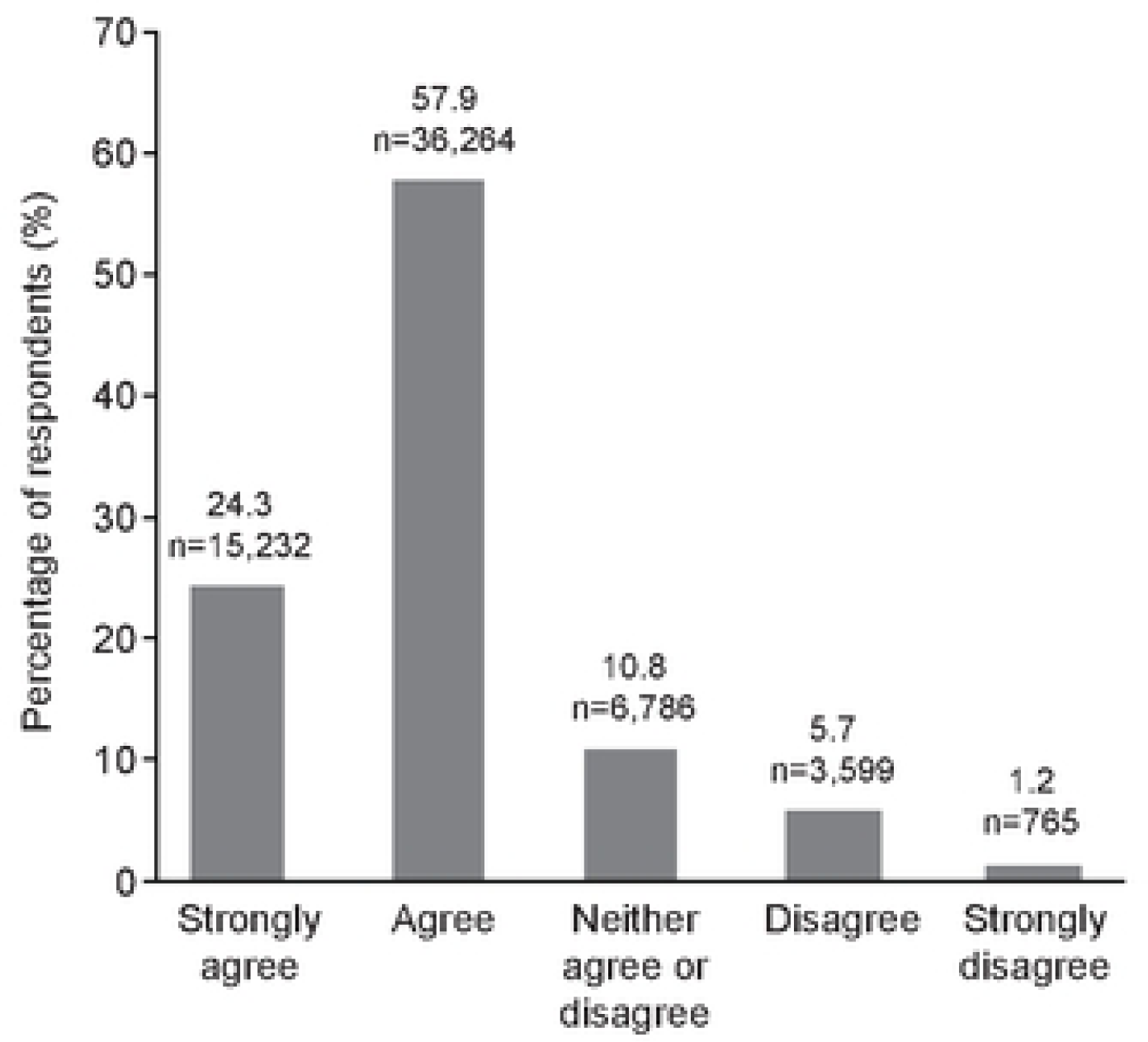
Likelihood that respondents will use the HMTK program as part of their mental health and wellbeing curriculum. Base n=62,646. **Q. How likely are you to use the videos from this program in your classroom?**

**Fig 6.**
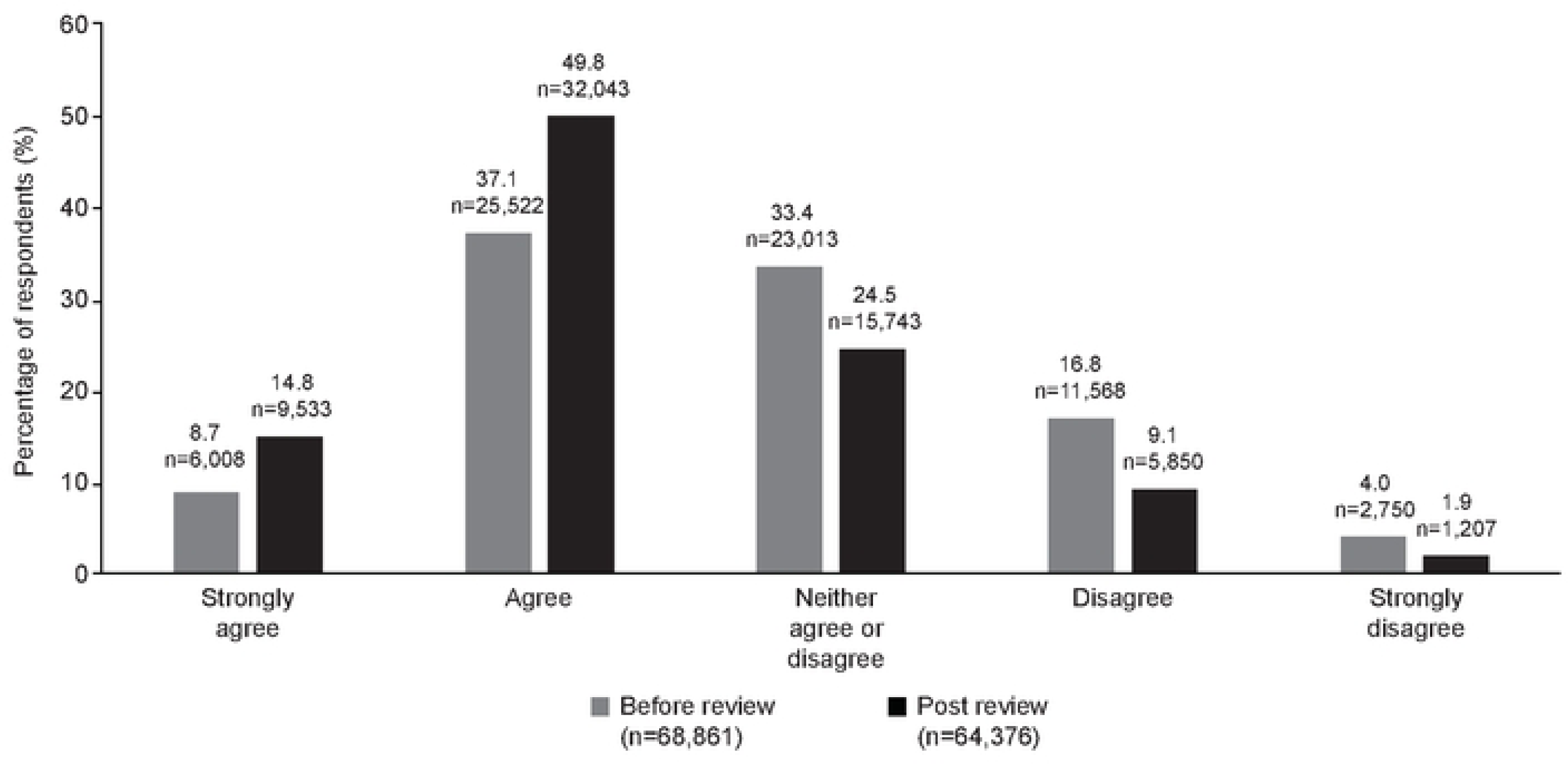
Perceptions of support of educators by the State of California before review of the HMTK program and following review and download of the HMTK program. **Pre-review n=68,861; post-review n=64,376** **Q. I feel the State of California is committed to supporting the social and emotional learning needs of teachers and students.**

Educators accessing the portal and watching at least 50% of the HMTK video program were eligible for a financial reward of $100. When educators were asked following their review of the program what would motivate them to use it in their classroom, 34.1% said that integrating the program into their own mental health and wellbeing curriculum was the motivating factor, with 32.3% indicating that financial inducements would incentivize the program’s use; results were comparable across all student cohorts (**Fig 7**).

**Fig 7.**
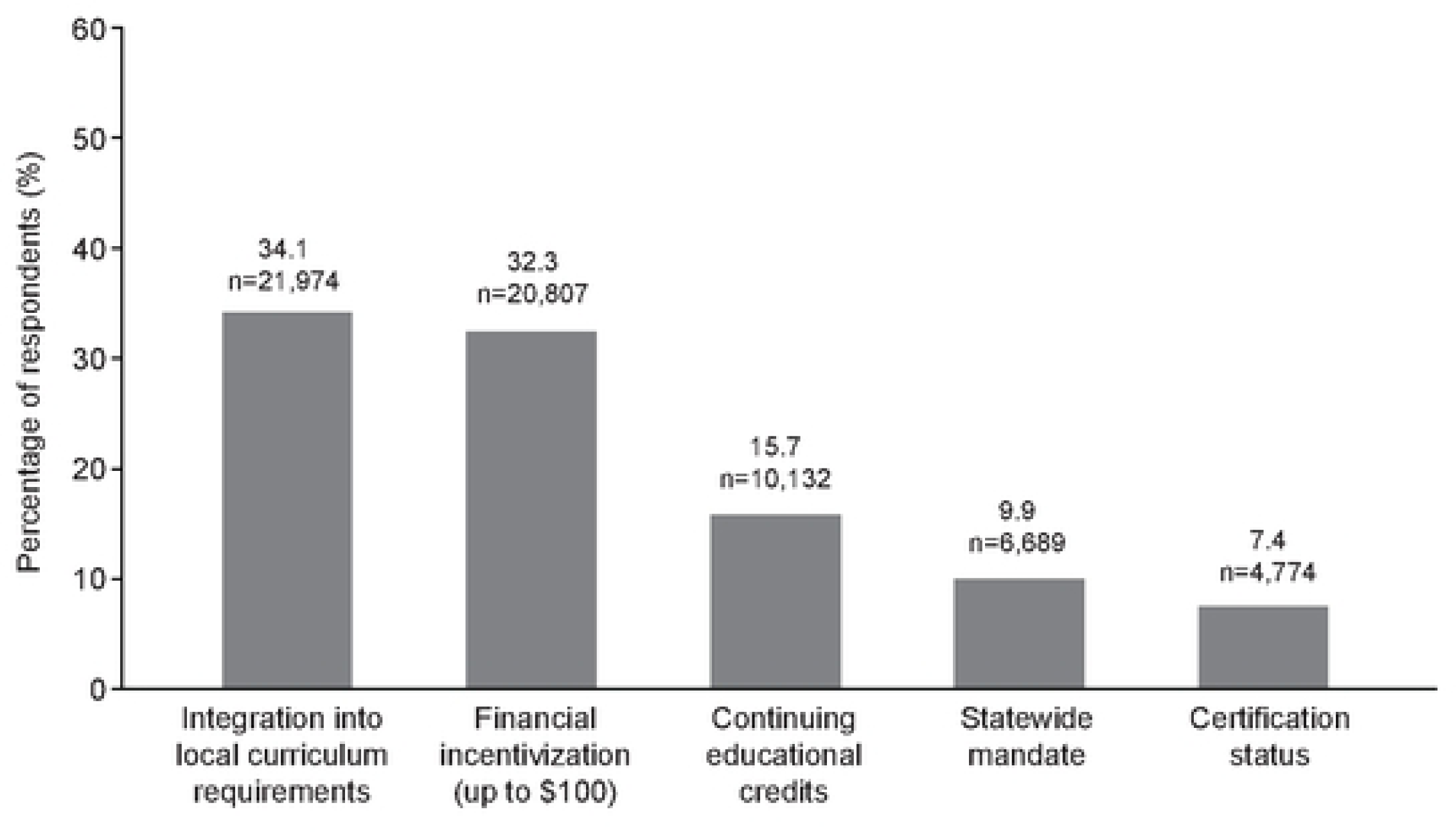
Respondents’ rationale for integrating the HMTK program into their curriculums supporting mental health and wellbeing. Base n=64376. **Q. What would best motivate teachers like you to use the videos from this program in your classroom?**

## Discussion

This survey of more than 60,000 educators provides insight into the impact of the COVID-19 pandemic on young people and the needs of educators to build robust and engaging mental health and wellbeing promotion programs. School is an ideal and critical setting to promote and support good mental health and wellbeing for young people. Schools have been flagged consistently as the most appropriate setting by policymakers, providing a common setting for all young people regardless of ethnicity or socioeconomic status. [24–26]

Evidence-based mental health prevention programs have demonstrated efficacy in terms of emotional and behavioral problems. [27] Given the evidence indicating the early emergence of mental health and learning disorders during childhood and adolescence, prevention programs focused on building mental health skills among school age young people are incredibly valuable in decreasing the severity of symptom onset while also providing children and teens with the skills they need to cope with stress. [6]

Educators recognize that student mental health and wellbeing is a central part of their students’ readiness to learn, but educators can often feel overwhelmed by the complexity and volume of the issues they are confronted with. They may also feel they lack mental health literacy necessary to address mental health and wellbeing effectively. [28–30] Data from this survey similarly suggests that almost all educators need support materials to help them promote student mental health and wellbeing. However, given the timing of this survey, it cannot be known how the need for support and resources for educators differs from pre-pandemic needs or if an enhanced need for mental health and wellbeing interventions will continue as young people continue to cope with the evolution of the pandemic. This survey has identified a high need among educators for tools in developing learning resources for their students, and survey responses clearly indicate the need for these resources to be engaging and appropriate for the age range they teach. Up to a quarter of respondents suggested that currently available materials were not particularly engaging, especially for older students.

This survey suggests that the HMTK video program is a valuable and complementary resource to school curricula to improve the mental health and wellbeing of young people. More than 80% of respondents endorsed that they had acquired useful insights from the program that they can take forward into the classroom. Almost all respondents believed the HMTK program would benefit their students and that their students would find these materials interesting and engaging. Respondents demonstrated a high willingness to implement the video program with their students. Encouragingly, the impact of the HMTK video materials was comparable across all educators regardless of the age range of their students, suggesting that the differentiated content of the HMTK program addresses the specific needs of each age group effectively.

The data from the HMTK portal suggests that the need to promote and sustain mental health and wellbeing is comparable in all age groups of young people, which is perhaps surprising given that it might be expected that the impact of COVID-19 lockdown might affect age groups differently. However, data exploring the impact of COVID-19 on younger children is lacking compared with that for adolescents. [10, 18] Studies of social and emotional prevention programs such as mindfulness have demonstrated promise [31], but findings are mixed [32], and further insights are required. It may be that prevention programs need to be more holistic and grounded in a range of evidence-based concepts, moving beyond just mindfulness to explore elements of emotion management, relaxation, and understanding thinking patterns. A broader approach helps to build mental health literacy and coping skills among young people accessing the program, and this approach may help to circumvent the effects of individual differences associated with only a narrow range of skills taught.

Building mental health literacy among educators is important to equip them to provide effective student support. However, implementation of preventative mental health and wellbeing programs is time consuming and needs to compete with a range of other priorities for educators, along with often limited training and access to resources. [28] It is interesting to note that whilst financial inducements were considered the most powerful incentive by almost a third of respondents, the potential to use the HMTK video program to enhance their teaching because they were engaging was equally motivating. Only a small percentage (10.4% overall, 11.5% K–5, 14.0% Grades 6–8, and 8.2% in Grades 9–12) said they would implement the HMTK program only if mandated to, suggesting that respondents recognize the critical need for these materials and value their potential in enhancing students’ academic engagement.

Whilst the survey cohort was robust, these data are limited by several factors, particularly with regard to the fact that survey questions were only able to track self-reported implementation or intent to implement rather than surveying actual rates of implementation and outcomes on young people. Similarly, these data only capture feedback about the program from the perspective of educators. Understanding the impact of the HMTK program from the perspective of students is an imperative and is the subject of ongoing study. Likewise, understanding the impact of the program once it is more widely implemented across more States will be evaluated further with additional studies. It would also be compelling in future study to understand the impact on students who view the program and its videos against a control group who do not. Whilst data are lacking in the research literature about expectations for implementation and uptake rates of mental health and wellbeing programs in school, it is clear that implementation of school-based social and emotional learning (SEL) support is often a multi-faceted, multi-year process with many factors involved affecting educators’ ability to adopt and utilize resources with students.

An unexpected element of this survey was the increase in educators’ belief that the State of California is committed to supporting the social and emotional learning of their students. Before review of the HMTK program, 45% of respondents believed that the State of California was committed to supporting them in delivering mental health and wellbeing promotion programs. Post-review of the HMTK program, more than 64% of educators believed the State of California was committed to supporting this need, and the percentage of respondents stating a lack of commitment reduced by half. This suggests that sponsorship of programs like HMTK at the State level can shift public perception and directly empower educators to enhance their support for the mental health and wellbeing of their students.

The strength of this survey is its high number of respondents, providing insight into issues facing educators across California and the challenges young people face in their daily lives. The HMTK video program and its positive reception by educators suggest that these resources can play a critical role in supporting young people, opening up conversations about mental health and wellbeing and helping K-12 students to navigate the mental health stressors that they encounter on the path to adulthood. A companion survey exploring the impact of the complete HMTK program on educators and its potential implementation in the classroom is underway. These data will provide even more granular insight into the potential for the HMTK program across California.

In conclusion, there is growing evidence of a positive effect on the mental health of children and adolescents via active implementation of preventative mental health and wellbeing programs. These programs can play a valuable role in preventing onset or severity of mental health symptoms and promoting emotional wellbeing. This survey has demonstrated the potential usefulness of a USA-specific prevention program for use across a wide range of educational settings. Studies demonstrating the impact of this program on the mental health and wellbeing of young people are underway, and data demonstrating its impact are eagerly awaited.

## Data Availability

All data are provided in the supporting information

## Acknowledgements

The authors are grateful to Lake Research Partners for assistance in developing the survey and PDG for its implementation. Editorialassistance and support was provided by Susan Allen, Brandfish Limited UK.

